# What interventions or best practice are there to support people with Long COVID, or similar post-viral conditions or conditions characterised by fatigue, to return to normal activities: a rapid review

**DOI:** 10.1101/2023.01.24.23284947

**Authors:** Llinos Haf Spencer, Annie Hendry, Abraham Makanjuola, Bethany F Anthony, Jacob Davies, Kalpa Pisavadia, Dyfrig Hughes, Deb Fitzsimmons, Clare Wilkinson, Rhiannon Tudor Edwards, Ruth Lewis, Alison Cooper, Adrian Edwards

## Abstract

Previous research has categorised symptoms of COVID-19 / Long COVID into 12 thematic areas including: fever, myalgia, fatigue, impaired cognitive function, and that COVID-19 survivors had reduced levels of physical function, activities of daily living, and health-related quality of life. Our aim was to review the evidence for interventions or best practice to support people with Long COVID, or similar post-viral conditions characterised by fatigue, to return to normal activities.

Evidence was included from guidelines, systematic reviews (SR), and primary studies. The primary studies focussed on Long COVID (LC) indicated that there should be a needs-based focus to care for those with LC. Consideration should be given to individuals living with LC in the same way as people with disabilities are accommodated in terms of workplace adjustment. Two SRs indicated that non-pharmaceutical interventions (NPIs) for patients with LC or chronic fatigue syndrome could help improve function for activities of daily life. However, the third, most recent SR, concluded that there is a lack of robust evidence for NPIs. LC fatigue management methods may be beneficial under certain conditions. One SR reported work capability as an outcome however they did not find any studies which evaluated the impact of interventions on return to work/ normal life. One primary study, on individuals with CFS, described a written self-management programme. Following this intervention there was an 18% increase in the number of patients in employment.

Policy and practice implications: Long COVID is still being established as a post-viral condition with many symptoms. Patient-centred treatment options such as occupational therapy, self-management therapy and talking therapy may be considered in the same way as for other debilitating conditions. Return-to-work accommodations are needed for all workers unable to return to full-time employment. Due to the nature of the studies included, there was little reported evidence of effectiveness of getting individuals back into their normal activities.

**Funding statement:** The Bangor Institute for Health and Medical Research was funded for this work by the Wales COVID-19 Evidence Centre, itself funded by Health & Care Research Wales on behalf of Welsh Government.

**Rapid Review Details:** *Review conducted by:* Bangor Institute for Health and Medical Research (BIHMR), Bangor University.

*Review Team:*

▪ Dr Llinos Haf Spencer,
▪ Dr Annie Hendry,
▪ Mr Abraham Makanjuola,
▪ Ms Bethany Fern Anthony,
▪ Mr Jacob Davies,
▪ Ms Kalpa Pisavadia,
▪ Professor Dyfrig Hughes,
▪ Professor Deb Fitzsimmons,
▪ Professor Clare Wilkinson,
▪ Professor Rhiannon Tudor Edwards

*Review submitted to the WCEC on:* 11 January 2023

*Stakeholder consultation meeting:* 8^th^ November 2022

*Rapid Review report issued by the WCEC in:* January 2022

*WCEC Team:* Adrian Edwards, Ruth Lewis, Alison Cooper and Micaela Gal involved in drafting the Topline Summary and editing.

*This review should be cited as:* RR00042_ Wales COVID-19 Evidence Centre

*Disclaimer:* The views expressed in this publication are those of the authors, not necessarily Health and Care Research Wales. The WCEC and authors of this work declare that they have no conflict of interest.

*TOPLINE SUMMARY:* What is a Rapid Review?
Our rapid reviews (RR) use a variation of the systematic review (SR) approach, abbreviating or omitting some components to generate the evidence to inform stakeholders promptly whilst maintaining attention to bias. They follow the methodological recommendations and minimum standards for conducting and reporting RR, including a structured protocol, systematic search, screening, data extraction, critical appraisal, and evidence synthesis to answer a specific question and identify key research gaps. They take 1 to 2 months, depending on the breadth and complexity of the research topic/question(s), extent of the evidence base, and type of analysis required for synthesis. Who is this summary for?
Policymakers in Welsh Government to plan and deliver services for individuals with Long COVID as they re-enter training, education, employment, and informal caring responsibilities. Background / Aim of Rapid Review
Previous research has categorised symptoms of COVID-19/Long COVID into 12 thematic areas including: fever, myalgia, fatigue, impaired cognitive function, and that COVID-19 survivors had reduced levels of physical function, activities of daily living, and health-related quality of life (Amdal et al., 2021; de Oliveira Almeida et al., 2022). NICE guidelines highlight the impact of the condition on quality of life and the challenge of determining best practice based on the current evidence (National Institute for Health and Care Excellence et al., 2022). Treatments for other post-viral syndromes may also apply to people living with Long COVID (Wong and Weitzer, 2021). Our aim was to review the evidence for interventions or best practice to support people with Long COVID, or similar post-viral conditions characterised by fatigue, to return to normal activities (including return to the workforce, education, childcare, or housework). Key Findings
Evidence was included from guidelines (n=3), systematic reviews (SRs) (n=3), and primary studies (n=4). Extent of the evidence base

▪ Two SRs included non-pharmacological interventions for Long COVID or post-viral syndromes, including Long COVID (Chandan et al., 2022; Fowler-Davis et al., 2021). The remaining SR focused on interventions for Chronic Fatigue Syndrome (CFS).
▪ The four primary studies were conducted in the UK, USA, Norway, and Turkey. The SRs included studies from across Europe, Asia, Africa, and Australasia.
▪ Included SRs and primary studies evaluated non-pharmaceutical interventions, including fatigue management, exercise therapy, Cognitive Behavioural Therapy (CBT), workplace support, self-management, sleep therapy, music therapy, and counselling.
▪ Two relevant guidelines were identified for Long COVID and one for ME/CFS. The Long COVID guideline was aimed at employers, and the ME/CFS guideline was aimed at service providers and users. Recency of the evidence base

▪ Included papers were from 2014 to 2022. Evidence of effectiveness

▪ The primary studies focussed on Long COVID indicated that there should be a needs-based focus to care for those with Long COVID (Lunt et al., 2022; Skilbeck, 2022; Wong et al., 2022). Consideration should be given to individuals living with Long COVID in the same way as people with disabilities are accommodated in terms of workplace adjustment (e.g. part-time hours, working from home, or hybrid working).
▪ Two SRs indicated that non-pharmaceutical interventions for patients with Long COVID or CFS could help improve function for activities of daily life (Fowler-Davis et al., 2021; Larun et al., 2019). However, the third and most recent SR concluded that there is a lack of robust evidence for non-pharmaceutical interventions (Chandan et al., 2022).
▪ Long COVID fatigue management by exercise therapy, electrical nerve stimulation, sleep and touch therapy, and behavioural self-management may be beneficial when: physical and psychological support is delivered in groups, people can plan their functional response to fatigue, strengthening rather than endurance is used to prevent deconditioning, fatigue is regarded in the context of an individual’s lifestyle and home-based activities are used (Fowler-Davis et al 2021).
▪ One SR (Chandan et al 2022) reported work capability as an outcome however they did not find any studies which evaluated the impact of interventions on return to work/ normal life.
▪ One primary study concentrated on individuals with CFS (Nyland et al., 2014). Nyland et al. (2014) described a written self-management programme featuring active coping (with CFS) strategies for daily life. Following this intervention, there was an 18% increase in the number of patients in employment (from baseline to follow-up) (Nyland et al., 2014). Best quality evidence

▪ The three SRs (Chandan et al., 2022; Fowler-Davis et al., 2021; Larun et al., 2019) were of high quality, as was one of the cohort studies (Lunt et al., 2022). Policy Implications

▪ Long COVID is still being established as a post-viral condition with many symptoms. The Welsh Government may seek to consider patient-centred treatment options such as occupational therapy, self-management therapy and talking therapy (such as Cognitive Behavioural Therapy) in the same way as for other debilitating conditions including ME/CFS.
▪ Return-to-work accommodations are needed for all workers unable to return to full-time employment.
▪ Due to the nature of the studies included, there was little reported evidence of effectiveness of getting individuals back into their normal activities. Strength of Evidence
Confidence in the findings is low. Only four primary studies reported outcomes relating to work capacity and return to normal activities such as childcare and housework.

## 1. BACKGROUND

### 1.1 Who is this review for?

Welsh Government stakeholders, including patient and public representatives, requested the Wales Covid Evidence Centre (WCEC) and the Bangor Institute for Health and Medical Research (BIHMR) team from Bangor University to look at interventions which may help individuals who have a diagnosis of Long COVID-19 (Kenny et al., 2022; National Institute for Health and Care Excellence et al., 2022) (and similar post-viral syndromes) to return to normal activities of life such as education, training, employment, and informal caring responsibilities. The interventions may include self-management, rehabilitation, exercise, or any other intervention related to helping individuals return to the activities the individual had undertaken prior to COVID-19 disease.

#### Background / Aim of Rapid Review

In a Systematic Review (SR) published in 2021 (Amdal et al., 2021), 75 distinct symptoms of COVID-19/Long COVID were categorised into 12 thematic areas: from general symptoms such as fever, myalgia, and fatigue, to neurological and psychological issues. The SR revealed three extra issues experienced during active disease and long-term problems: fatigue, psychological issues, and impaired cognitive function. Another SR focussing on the survivors of COVID-19 reported that survivors had reduced levels of physical function, activities of daily living, and health-related quality of life (de Oliveira Almeida et al., 2022). Furthermore, incomplete recovery of physical function and performance in activities of daily living were observed 1 to 6 months post-infection (de Oliveira Almeida et al., 2022).

##### NICE guidelines on Long COVID state that

*“This new and emerging condition, which has been described using a variety of terms including ‘Long COVID’, can have a significant effect on people’s quality of life. It also presents many challenges when trying to determine the best-practice standards of care based on the current evidence. There is no clinical definition or clear treatment pathway, and there is a minimal, though evolving, evidence base”* (National Institute for Health and Care Excellence et al., 2022, *p*.*3)*

#### Is Long COVID similar to Myalgic Encephalomyelitis (or encephalopathy) / Chronic Fatigue Syndrome (ME/CFS)?

One SR (Wong and Weitzer, 2021) outlined in the preliminary review of the evidence suggested that treatments for conditions such as Fibromyalgia, ME/CFS and other post-viral conditions may also apply to people living with Long COVID. This SR found that the symptoms of these conditions seem to be closely associated with Long COVID (e.g. prolonged symptom duration, fatigue, reduced daily activity and post-exertional malaise, neurological pain, memory and attention difficulties, thermostatic instability, dizziness and chest palpations/pain) (Wong & Weitzer, 2021).

#### Return to work as an example of returning to normal activities

The topic of work and keeping ‘active’ and ‘engaging’ (either through paid employment or unpaid work such as informal caring) is of importance as the Welsh and UK Governments want Prosperity for All (Welsh Government, 2017a, 2017b) and Equality for all (UK Legislation, 2010). Getting people into, or back into, education, training and employment are of high priority as being active and contributing to society are important for well-being and the economy (Edwards et al., 2019). In June 2022, the Welsh Government launched their £5 million support package for the new ‘Adferiad/Recovery’ programme to expand the provision of diagnosis, treatment and rehabilitation and care for those with Long COVID in Wales (Welsh Government, 2021). The NHS and National Institute for Health and Care Excellence (NICE) Guidelines for myalgic encephalomyelitis (or encephalopathy) / chronic fatigue syndrome (ME/CFS) highlight the importance of maintaining independence through an individual care and support plan (National Institute for Health and Care Excellence, 2021; NHS Plus, 2006). A recent review published in 2019 (Castro-Marrero et al., 2019) surmised that around 50% of ME/CFS patients do not work due to temporary or permanent disability, and previous authors have suggested that this figure could be higher at around 54% (Ross et al., 2004).

In terms of Long COVID, an SR including 11 studies on the impact of Long COVID on return to work was conducted in Italy (Gualano et al., 2022). The authors noted that there is a highly variable return to work rate after COVID-19, ranging from 10% to 100% and attributed some of the variances to healthcare systems, working cultures and habits in different countries. The 100% return to work rates were observed in China. No interventions or best practices to get people back into work were described in these papers.

### 1.2 Purpose of this review

The rapid review presented here is for policymakers in Welsh Government to plan and deliver services for those with Long COVID or similar post-viral conditions characterised by fatigue as they re-enter training, education, employment, and informal caring responsibilities after a period of ill health. The research question is as follows: ‘What interventions or best practice is there to support people with Long COVID, or similar post-viral conditions characterised by fatigue to return to normal activities?’ The review is focused on outcomes relating to work capacity or ability to conduct ‘normal life activities’ (such as childcare or housework).

## 2. RESULTS

### 2.1 Overview of the Evidence Base

Ten relevant studies published between 2014 to 2022 were included in this rapid review (see Tables 2.1 and 2.2). The evidence base comprised of clinical guidelines (n=3) systematic reviews (n=3), cohort studies (n=2), a qualitative study (n=1), and a case study (n=1). In general, this evidence focused on exercise therapy, CBT, self-management therapy, and counselling. However, two SRs also included music therapy, tele-rehabilitation, neuro modulation and a range of interventions such as acupuncture, homeopathy, yoga, electrical nerve stimulation, sleep therapy, touch therapy and psycho-spiritual education (Chandan et al., 2022; Fowler-Davis et al., 2021). There was no evidence of interventions relating to return to work or returning to training, suggesting an evidence gap in these areas.

The authors of this rapid review are also aware of six further ongoing UK NIHR-funded non-pharmacological studies on Long COVID, which are due to be finalised in January, February, August, and October 2023 (See Appendix 3).

Two SRs included non-pharmacological interventions for Long COVID or post-viral syndromes, including Long COVID (Chandan et al., 2022; Fowler-Davis et al., 2021). The remaining SR focused on interventions for CFS (Larun et al., 2019). All three SRs were of high quality.

Only one of the primary studies was of high quality (Lunt et al., 2022). This study had a focus on Long COVID and suggested workplace accommodations for workers returning to work after a Long COVID diagnosis (Lunt et al., 2022). The other primary studies, with moderate quality ratings, focused on needs-based approaches for returning to normal function after a Long COVID diagnosis. These studies suggest that treatments such as management therapy and counselling as rehabilitation interventions may benefit individuals with Long COVID (Skilbeck, 2022; Wong et al., 2022).

The Nyland et al (2014) paper, although focusing on CFS and not Long COVID, provided some evidence on self-management to support young people with CFS to gain employment (Nyland et al., 2014). This was the only included report of this kind found in the review searches, which included information relevant to this rapid review.

#### 2.1.1 Guidelines

Three guidelines were identified in relation to Long COVID or ME/CFS. Two were related to COVID-19, one from the Chartered Institute for Personnel and Development (Chartered Institute of Personnel and Development, 2022; National Institute for Health and Care Excellence, 2021) and the other from the National Institute for Health and Care Excellence (NICE; 2022). The third identified guideline was also from NICE and focused on ME/CFS (National Institute for Health and Care Excellence, 2021).

The CIPD (Chartered Institute of Personnel and Development, 2022) emphasise the importance of ensuring full access to workplace adjustments to enable individuals with Long COVID to sustain work. The CIPD stated that as the symptoms of Long COVID are likely to fluctuate over time, different work adjustments may be needed at different times. Therefore, it is vital to have ongoing and supportive conversations to review work adjustments which may include home or hybrid working, other flexible working arrangements, reduced or off-peak commuting time, adapting work tasks to make them less physically, mentally, or cognitively demanding, and providing a wellbeing room for downtime during the working day.

The NICE (National Institute for Health and Care Excellence (NICE), 2022; National Institute for Health and Care Excellence, 2021) guidelines suggest that people with disabilities may need reasonable adjustments in the workplace, especially when there are other health and safety matters to consider.

The evidence review conducted for the NICE guideline on ME/CFS covered a wide range of non-pharmacological interventions to improve mobility and/or general wellbeing. The researchers acknowledged the controversy regarding the previous guidance, published in 2007, which recommended the use of CBT and Graded Exercise Therapy (GET) (National Institute for Health and Care Excellence, 2021). The recent guidance recommends that people experiencing fatigue ‘should undertake therapy options where they remain within their energy limits, and care should be given to undertake activities that do not worsen symptoms’ (National Institute for Health and Care Excellence, 2021).

#### 2.1.2 Systematic reviews

Three high quality SRs (Chandan et al., 2022; Fowler-Davis et al., 2021; Larun et al., 2019) reported on how post-viral syndrome and Long COVID populations can be helped to improve function for activities of daily life. Within the three reviews, 53 studies were included (Fowler-Davis et al., 2021, n=40; Larun et al., 2019, n=8; Chandan et al 2022, n=5).

The Fowler-Davis et al., (2021) SR suggested that Long COVID fatigue management by exercise therapy, electrical nerve stimulation, sleep and touch therapy, and behavioural self-management may be beneficial when:

- physical and psychological support is delivered in groups
- people can plan their functional response to fatigue
- strengthening rather than endurance is used to prevent deconditioning
- fatigue is regarded in the context of an individual’s lifestyle
- home-based activities are used

In the Larun et al., (2019) SR, exercise therapy was compared with other interventions for symptoms of CFS, including fatigue, pain, physical function, quality of life, mood disorders, sleep, and service use (Larun et al., 2019). The exercise therapy included: swimming, cycling, and walking compared to usual care. The authors concluded that exercise therapy may have a positive effect on fatigue in adults with CFS compared with usual care or passive therapies (including cognitive behavioural therapy (CBT), adaptive pacing, and antidepressants). However, the authors were uncertain about the long-term improvements and the risk of side effects. Therefore, due to limited evidence, it was difficult for the authors to draw strong conclusions as to the comparative effectiveness of CBT, adaptive pacing, or other interventions in treating CFS.

The third systematic review reported that there is a lack of evidence overall relating to post-viral syndromes and non-pharmaceutical interventions (Chandan et al., 2022). This review included five studies describing five distinct interventions to support people with post-viral syndromes (Chandan et al., 2022). Four of these five studies (evaluating tele-rehabilitation, resistance exercises, pilates, and neuromodulation) reported statistically significant benefits in their primary outcomes. However, the fifth study (of music therapy combined with CBT) did not report any significant improvement in their primary outcomes (Chandan et al., 2022). The study was not adequately powered; therefore, further work may be necessary to assess efficacy (Chandan et al., 2022).

#### 2.1.3 Primary studies

Three of the included primary studies focused on Long COVID (Lunt et al., 2022; Skilbeck, 2022; Wong et al., 2022) and one primary study concentrated on individuals with CFS (Nyland et al., 2014). See Table 2.

##### Long COVID primary papers

The Lunt et al., (2022) paper described a mixed-methods study, including a survey regarding returning to work following COVID-19 recovery in England. An exploratory online survey was used to collect information regarding return-to-work experiences and recommendations of the participants. They found that only 15% of workers managed a full return to work, and 90% had experienced at least one post-COVID symptom, most commonly fatigue and cognitive effects. The authors suggested that some barriers to returning to work included symptom unpredictability, unhelpful attitudes to Long COVID recovery, and unrealistic expectations of employee capacity. Practical solutions reported by the participants of this study included: prioritisation of workload by the manager; maintaining contact with the sick employee during their leave in a respectful manner; buddy systems and open discussions to create reasonable expectations; ‘pacing’; allowing employees more time to settle back into their roles, with no time limits in the return to work process; evaluation of sickness absence policies to mitigate against bullying and harassment, and Long COVID awareness programmes to raise awareness and raise compassion for Long COVID sufferers in the workplace (Lunt et al., 2022).

The case study on Long COVID based in England focused on self-management therapy as an intervention for adjusting to life after Long COVID (Skilbeck, 2022). Self-management included psychoeducation on how to manage and monitor symptoms whilst also considering fluctuations in symptoms. It also involved assisting individuals to use goal-setting methods and encouraged seeking support from family and friends. Functional impairment was assessed using the Work and Social Adjustment Scale (WSAS), a reliable and valid tool (Zahra et al., 2014). The male participant reported an improvement in his depression and anxiety symptoms following self-management therapy as an intervention. He also reported improved quality of life as well as work and social adjustment. The participant reported that accepting his symptoms and connecting with his values played a role in his process of therapy. He also reported that he had learnt to take ownership of his recovery and that patience and allowing time were also key factors.

A qualitative study from the USA described the results from two focus groups with counsellors and physicians who treated people living with Long COVID (Wong et al., 2022). They aimed to describe the challenges individuals with Long COVID encountered when returning to work. Although this qualitative work did not evaluate an intervention, the findings are useful in planning future interventions. The focus group interview results highlighted the return-to-work issues of individuals with Long COVID. Accommodations such as gradual return to work were frequently mentioned by the healthcare professionals in the focus group interviews:

> “There are certain jobs that will not accommodate the patients and say, “if you can’t do X, Y, and Z, you can’t come back to work.” (P.29) (Wong et al., 2022)

> “I have another client who was also depressed. He was a part of three different COVID support groups. One at [the outpatient centre]. And, then he had found two [other support groups] on his own that he participated in. He found that very helpful to talk to other people who were experiencing the same kinds of uncertainties” (P.30) (Wong et al., 2022)

In summary, the Long COVID papers indicated that there should be a needs-based focus to care for those with Long COVID (Lunt et al., 2022; Skilbeck, 2022; Wong et al., 2022). Consideration should be given to individuals living with Long COVID in the same way people with disabilities are accommodated in terms of workplace adjustment (e.g. part-time hours, working from home, or hybrid working).

##### Chronic Fatigue Syndrome primary paper

The Nyland et al. (2014) paper described a longitudinal cohort study conducted in Norway, focusing on a written self-management programme featuring active coping [with CFS] strategies for daily life. The participants were 111 young patients with CFS after mononucleosis. Baseline was between 1996 and 2006, and follow-up was in 2009. All the patients that were unemployed at baseline received some form of disability benefit. This financial support and engagement with the social welfare system encouraged rehabilitation activities directed at finding new employment whilst unemployed. Following the self-management programme intervention there was an 18% increase in the number of patients in employment from contact from baseline (10% in employment) to follow-up (28% in employment). Logistic regression analyses showed that being employed at follow-up was associated with lack of arthralgia (OR=0.3, p=0.028) and reporting improvement (OR=1.8, p=0.062) at baseline. Another logistic regression analysis showed that being employed at contact 2 was associated with low FSS score at contact 2 (OR=0.53, p<0.001), lack of arthralgia (OR=0.40, p=0.041) and lack of concentration problems (OR=0.32, p=0.064), but none of the other symptoms reported at follow-up. The self-written management programme featured graded activity planning, a strategy which encourages physical activity relative to the individuals’ limitations to reduce fluctuations in CFS symptoms.

#### 2.1.4 Bottom line summaries for the secondary and primary included studies Secondary included studies

The findings of one SR suggest that fatigue management treatments for Long COVID may be limited and self-rated fatigue scales are a limitation of the research body as a whole (Fowler-Davis et al., 2021). Data synthesis from Fowler-Davis et al (2021) suggests that fatigue management is potentially beneficial when interventions are delivered in groups, where strengthening is used rather than endurance and where fatigue is considered in relation to individual lifestyles and activities (Fowler-Davis et al., 2021). Only one SR (Chandan et al., 2022) reported work capability as an outcome however they did not find any studies which evaluated the impact of interventions on return to work/ normal life. There was no evidence of interventions relating to return to training or returning to caring, suggesting an evidence gap in these areas.

##### Primary included studies

The primary studies indicated that there should be a needs-based focus to care for individuals with Long COVID in the same way care is presented to others with debilitating conditions. The self-management programme featuring active coping with CFS strategies for daily life seems effective in helping young people with CFS into employment. There was no evidence regarding training or informal care.

**Table 2.1.**
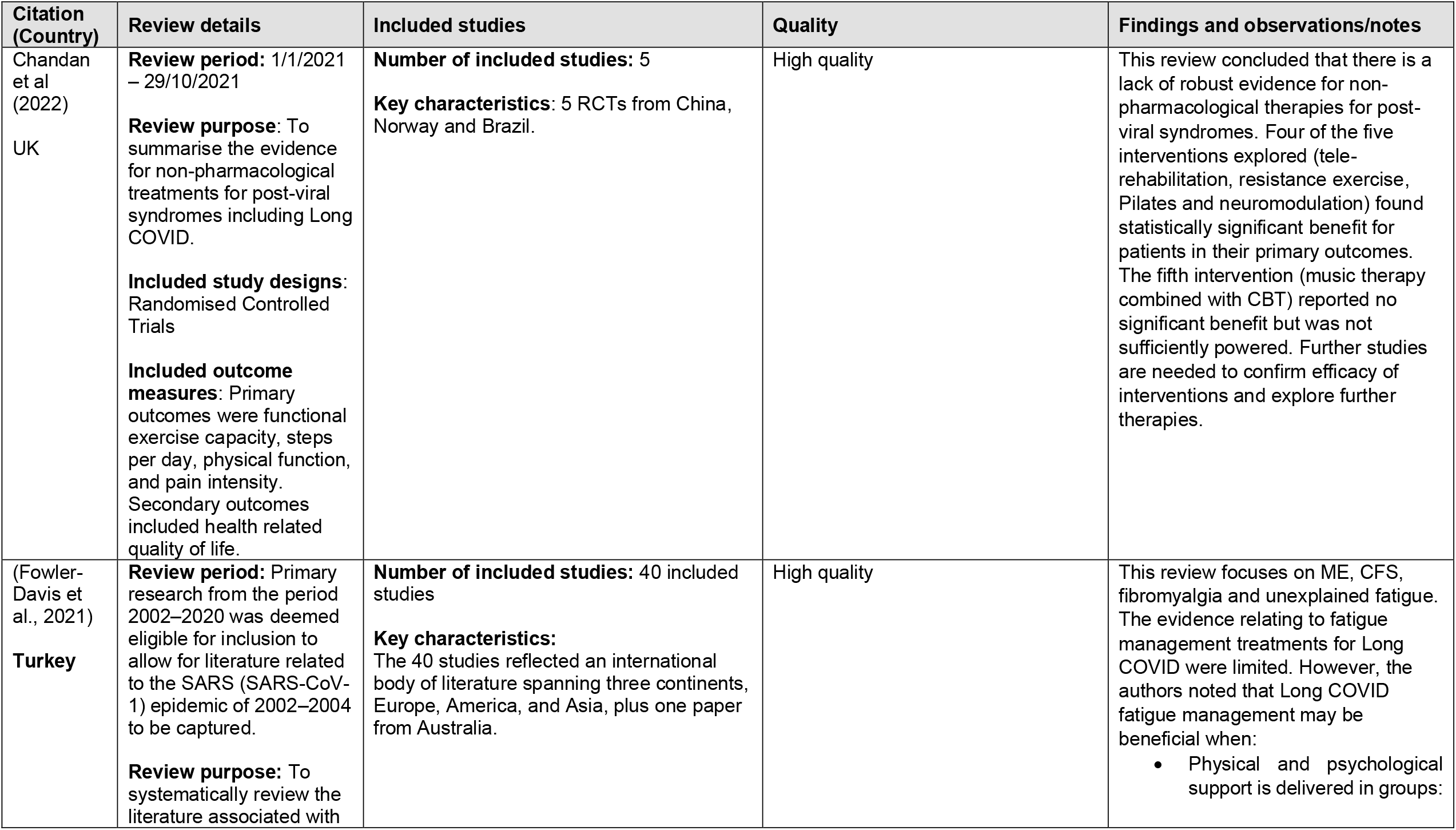
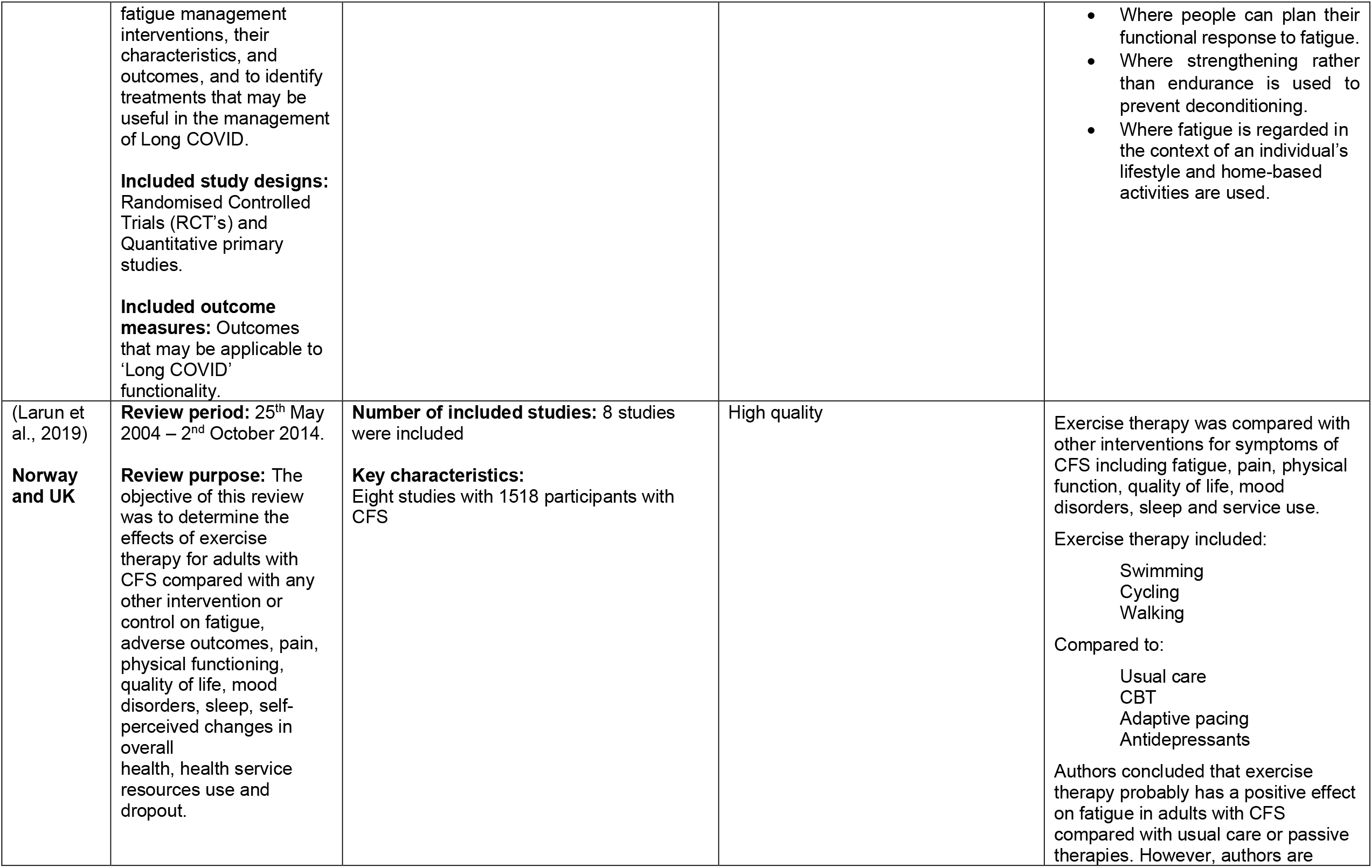
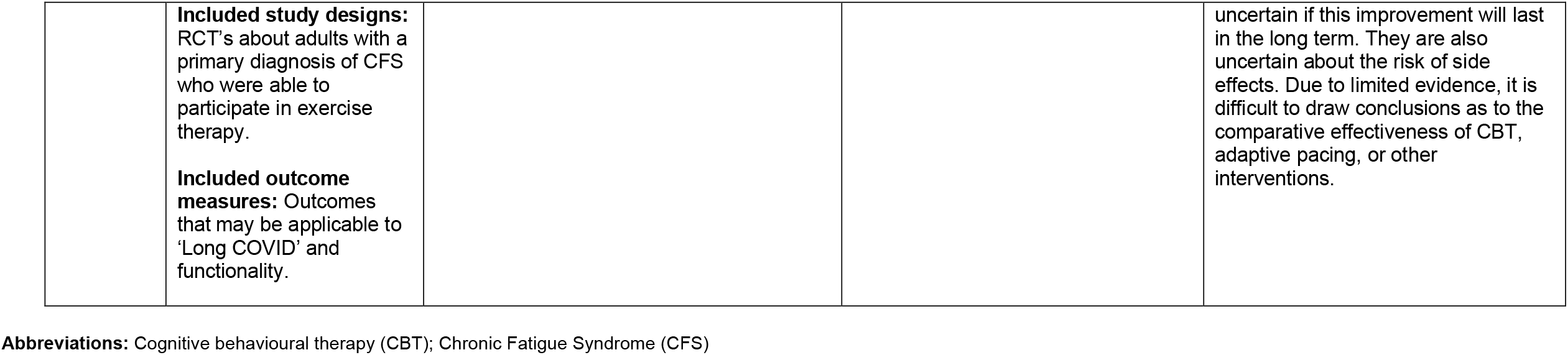
Summary of secondary research investigating Long COVID, or other conditions characterised by fatigue.

**Table 2.2.**
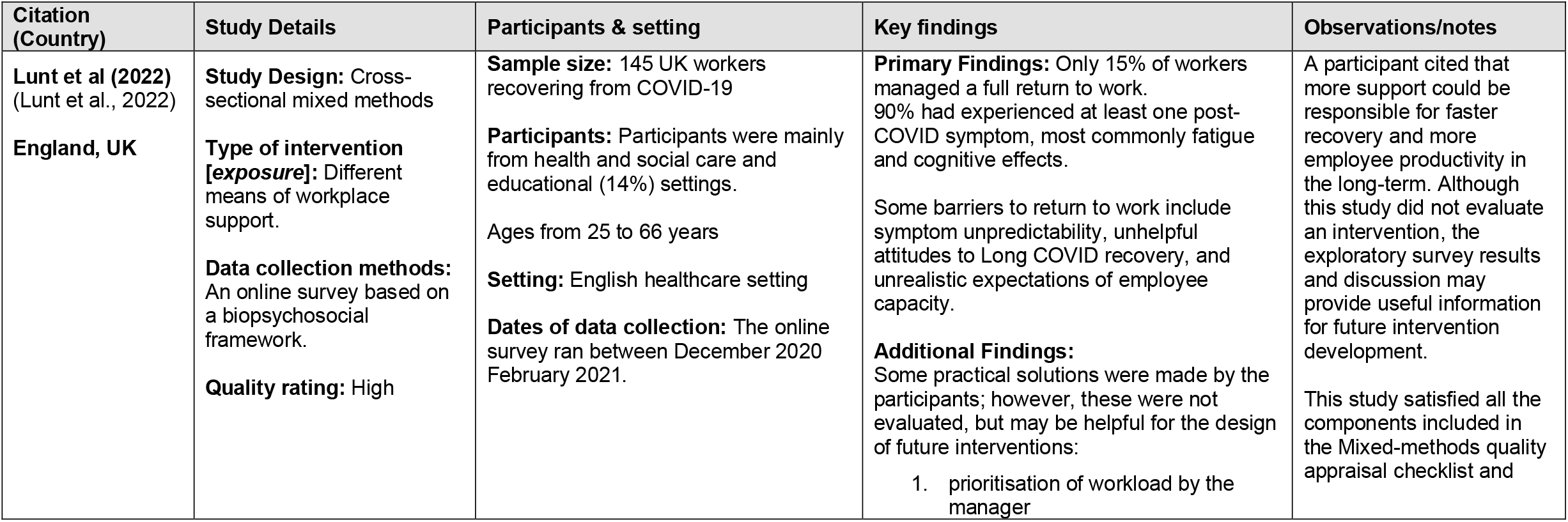
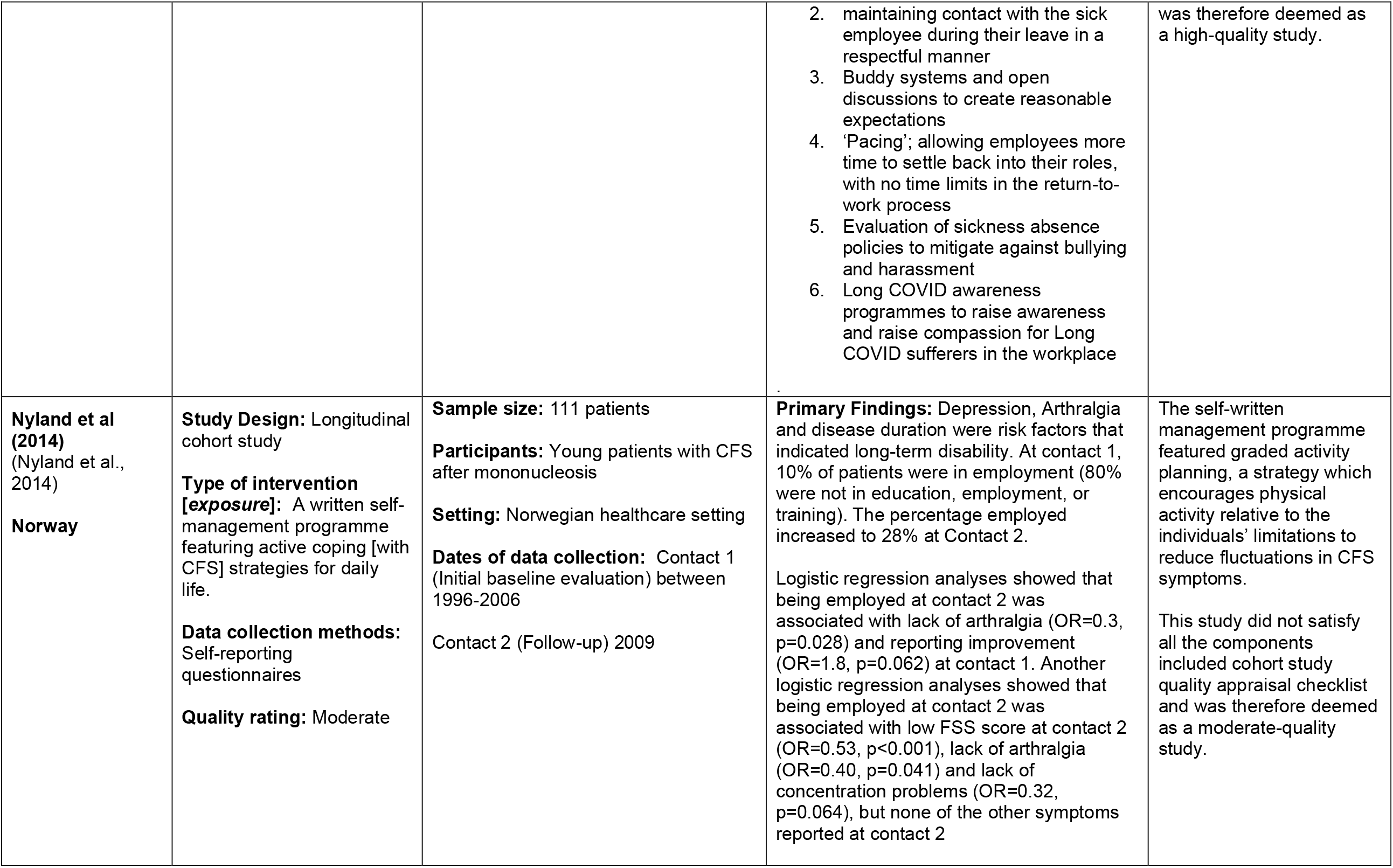
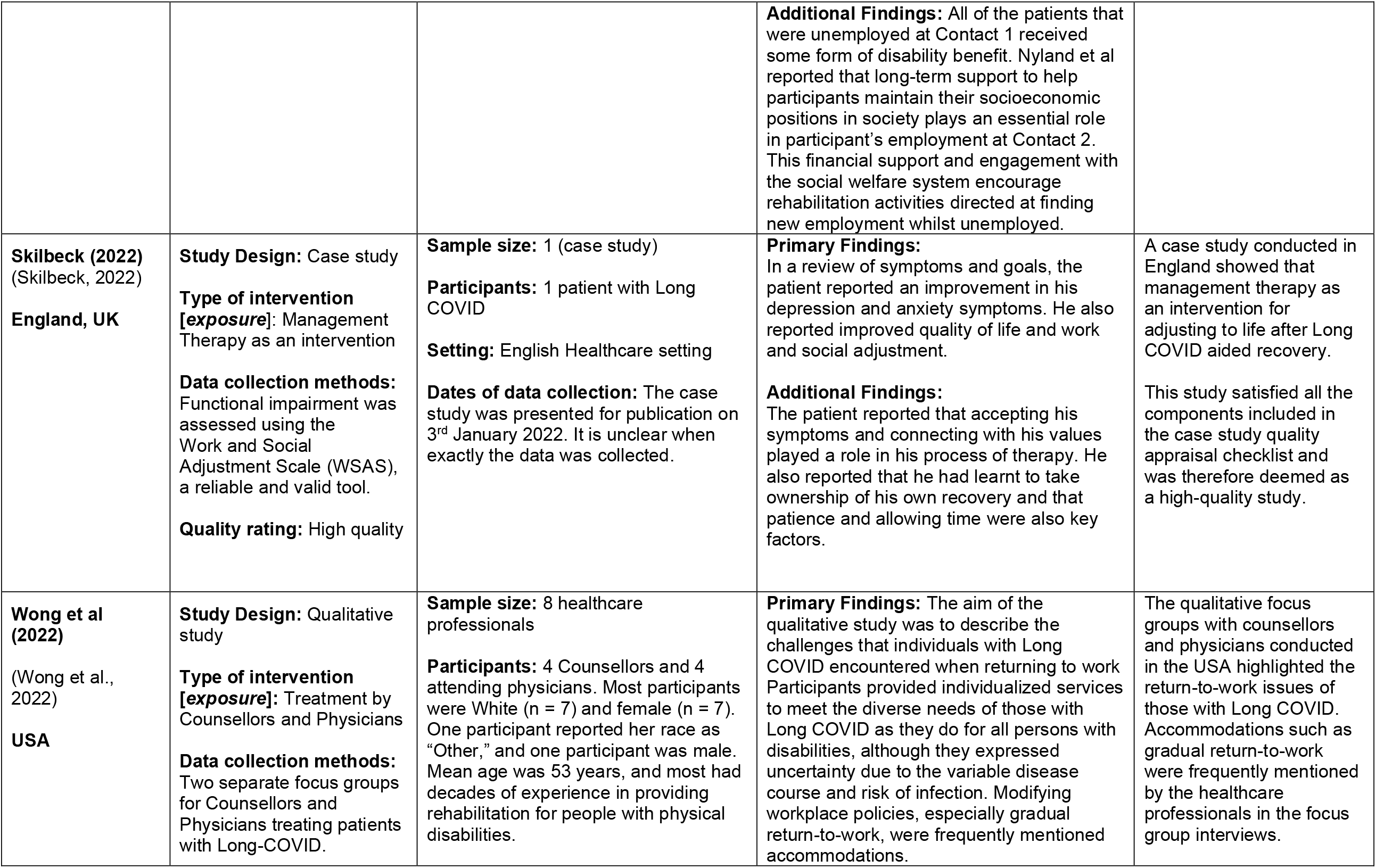
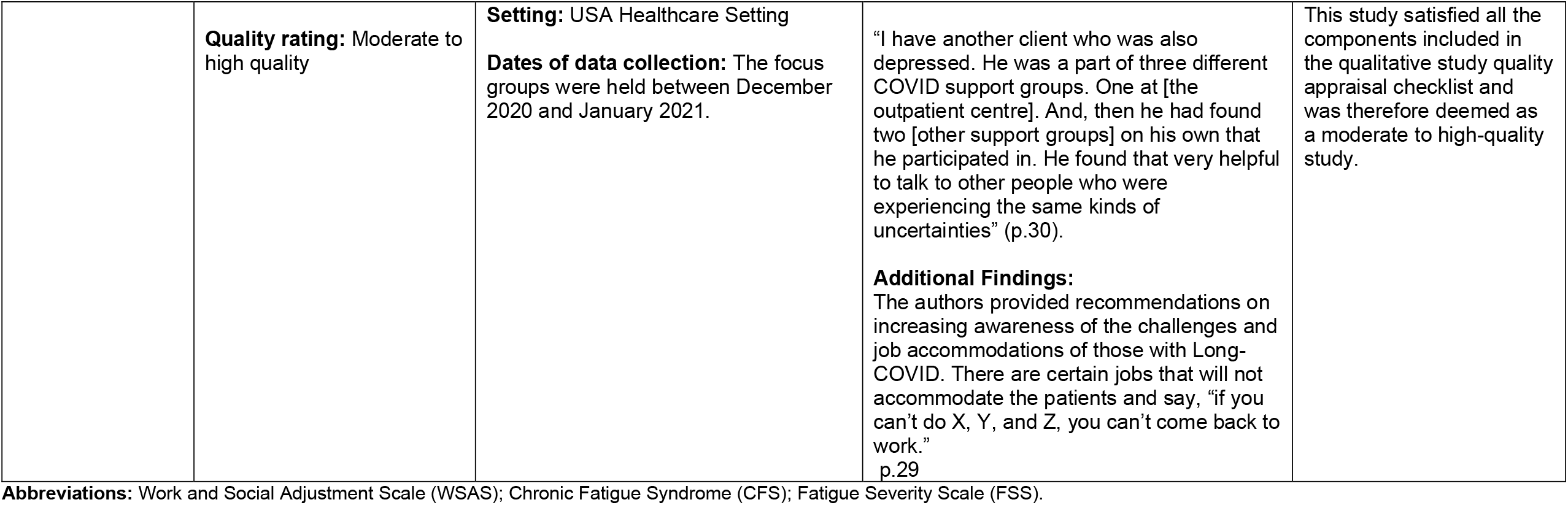
Summary of included primary evidence.

| Citation (Country) | Study Details | Participants & setting | Key findings | Observations/notes |
| --- | --- | --- | --- | --- |
| **Lunt et al (2022)**<br>(Lunt et al., 2022)<br>**England, UK** | **Study Design:** Cross-sectional mixed methods<br>**Type of intervention [exposure]:** Different means of workplace support.<br>**Data collection methods:** An online survey based on a biopsychosocial framework.<br>**Quality rating:** High | **Sample size:** 145 UK workers recovering from COVID-19<br>**Participants:** Participants were mainly from health and social care and educational (14%) settings.<br>Ages from 25 to 66 years<br>**Setting:** English healthcare setting<br>**Dates of data collection:** The online survey ran between December 2020 February 2021. | **Primary Findings:** Only 15% of workers managed a full return to work. 90% had experienced at least one post-COVID symptom, most commonly fatigue and cognitive effects.<br>Some barriers to return to work include symptom unpredictability, unhelpful attitudes to Long COVID recovery, and unrealistic expectations of employee capacity.<br>**Additional Findings:** Some practical solutions were made by the participants; however, these were not evaluated, but may be helpful for the design of future interventions:<br>1. prioritisation of workload by the manager | A participant cited that more support could be responsible for faster recovery and more employee productivity in the long-term. Although this study did not evaluate an intervention, the exploratory survey results and discussion may provide useful information for future intervention development.<br>This study satisfied all the components included in the Mixed-methods quality appraisal checklist and |
|  |  |  | maintaining contact with the sick employee during their leave in a respectful manner<br>Buddy systems and open discussions to create reasonable expectations<br>'Pacing'; allowing employees more time to settle back into their roles, with no time limits in the return-to-work process<br>Evaluation of sickness absence policies to mitigate against bullying and harassment<br>Long COVID awareness programmes to raise awareness and raise compassion for Long COVID sufferers in the workplace | was therefore deemed as a high-quality study. |
| **Nyland et al (2014)**<br>(Nyland et al., 2014)<br>**Norway** | **Study Design:** Longitudinal cohort study<br>**Type of intervention**<br>**[exposure]:** A written self-management programme featuring active coping [with CFS] strategies for daily life.<br>**Data collection methods:**<br>Self-reporting questionnaires<br>**Quality rating:** Moderate | **Sample size:** 111 patients<br>**Participants:** Young patients with CFS after mononucleosis<br>**Setting:** Norwegian healthcare setting<br>**Dates of data collection:** Contact 1 (Initial baseline evaluation) between 1996-2006<br>Contact 2 (Follow-up) 2009 | **Primary Findings:** Depression, Arthralgia and disease duration were risk factors that indicated long-term disability. At contact 1, 10% of patients were in employment (80% were not in education, employment, or training). The percentage employed increased to 28% at Contact 2.<br>Logistic regression analyses showed that being employed at contact 2 was associated with lack of arthralgia (OR=0.3, p=0.028) and reporting improvement (OR=1.8, p=0.062) at contact 1. Another logistic regression analyses showed that being employed at contact 2 was associated with low FSS score at contact 2 (OR=0.53, p<0.001), lack of arthralgia (OR=0.40, p=0.041) and lack of concentration problems (OR=0.32, p=0.064), but none of the other symptoms reported at contact 2 | The self-written management programme featured graded activity planning, a strategy which encourages physical activity relative to the individuals' limitations to reduce fluctuations in CFS symptoms.<br>This study did not satisfy all the components included cohort study quality appraisal checklist and was therefore deemed as a moderate-quality study. |
|  |  |  | **Additional Findings:** All of the patients that were unemployed at Contact 1 received some form of disability benefit. Nyland et al reported that long-term support to help participants maintain their socioeconomic positions in society plays an essential role in participant's employment at Contact 2. This financial support and engagement with the social welfare system encourage rehabilitation activities directed at finding new employment whilst unemployed. |  |
| **Skilbeck (2022)**<br>(Skilbeck, 2022)<br>**England, UK** | **Study Design:** Case study<br>**Type of intervention**<br>**[exposure]:** Management Therapy as an intervention<br>**Data collection methods:** Functional impairment was assessed using the Work and Social Adjustment Scale (WSAS), a reliable and valid tool.<br>**Quality rating:** High quality | **Sample size:** 1 (case study)<br>**Participants:** 1 patient with Long COVID<br>**Setting:** English Healthcare setting<br>**Dates of data collection:** The case study was presented for publication on 3<sup>rd</sup> January 2022. It is unclear when exactly the data was collected. | **Primary Findings:**<br>In a review of symptoms and goals, the patient reported an improvement in his depression and anxiety symptoms. He also reported improved quality of life and work and social adjustment.<br>**Additional Findings:**<br>The patient reported that accepting his symptoms and connecting with his values played a role in his process of therapy. He also reported that he had learnt to take ownership of his own recovery and that patience and allowing time were also key factors. | A case study conducted in England showed that management therapy as an intervention for adjusting to life after Long COVID aided recovery.<br>This study satisfied all the components included in the case study quality appraisal checklist and was therefore deemed as a high-quality study. |
| **Wong et al (2022)**<br>(Wong et al., 2022)<br>**USA** | **Study Design:** Qualitative study<br>**Type of intervention**<br>**[exposure]:** Treatment by Counsellors and Physicians<br>**Data collection methods:** Two separate focus groups for Counsellors and Physicians treating patients with Long-COVID. | **Sample size:** 8 healthcare professionals<br>**Participants:** 4 Counsellors and 4 attending physicians. Most participants were White (n = 7) and female (n = 7). One participant reported her race as "Other," and one participant was male. Mean age was 53 years, and most had decades of experience in providing rehabilitation for people with physical disabilities. | **Primary Findings:** The aim of the qualitative study was to describe the challenges that individuals with Long COVID encountered when returning to work. Participants provided individualized services to meet the diverse needs of those with Long COVID as they do for all persons with disabilities, although they expressed uncertainty due to the variable disease course and risk of infection. Modifying workplace policies, especially gradual return-to-work, were frequently mentioned accommodations. | The qualitative focus groups with counsellors and physicians conducted in the USA highlighted the return-to-work issues of those with Long COVID. Accommodations such as gradual return-to-work were frequently mentioned by the healthcare professionals in the focus group interviews. |
|  | **Quality rating:** Moderate to high quality | **Setting:** USA Healthcare Setting<br>**Dates of data collection:** The focus groups were held between December 2020 and January 2021. | “I have another client who was also depressed. He was a part of three different COVID support groups. One at [the outpatient centre]. And, then he had found two [other support groups] on his own that he participated in. He found that very helpful to talk to other people who were experiencing the same kinds of uncertainties” (p.30).<br>**Additional Findings:**<br>The authors provided recommendations on increasing awareness of the challenges and job accommodations of those with Long-COVID. There are certain jobs that will not accommodate the patients and say, “if you can’t do X, Y, and Z, you can’t come back to work.”<br>p.29 | This study satisfied all the components included in the qualitative study quality appraisal checklist and was therefore deemed as a moderate to high-quality study. |

**Abbreviations:** Work and Social Adjustment Scale (WSAS); Chronic Fatigue Syndrome (CFS); Fatigue Severity Scale (FSS).

## 3. DISCUSSION

### 3.1 Summary of the findings

The key evidence identified for this RR included guidelines (n=3), primary studies (n=4) and reviews (n=4) published since 2014 focusing on non-pharmacological interventions following either a diagnosis of Long COVID or ME/CFS. Primary studies focus on interventions to help people with post viral syndromes return to normal routines. The quality of the studies was appraised with appropriate quality appraisal tools according to their study design. Consequently, it was difficult to compare the quality of the evidence across the included studies.

#### 3.1.1 Summary of the guidelines

Three relevant guidelines were included. One of the guidelines referred specifically to the treatment of individuals living with ME/CFS (National Institute for Health and Care Excellence, 2021). The other two guidelines referred to Long COVID (Chartered Institute of Personnel and Development, 2022; National Institute for Health and Care Excellence et al., 2022). The guideline by CIPD was specifically related to working with symptoms of Long COVID, and the NICE Long COVID guidance was more general, with a focus on symptom management more so than return to normal activities of life.

#### 3.1.2 Summary of the systematic review evidence

Within the three systematic reviews included in this rapid review, 53 primary studies were included (Fowler-Davis et al., 2021, n=40; Larun et al., 2019, n=8; Chandan et al 2022, n=5). There is a lack of evidence for non-pharmacological interventions for post-viral syndromes (including Long COVID) and returning to activities of normal life or work (Chandan et al 2022). Larun et al (2019) conclude that there is uncertainty regarding the potential for long-term improvement and the possibility of side effects (Larun et al., 2019). Therefore, due to limited evidence, it is difficult to draw firm conclusions as to comparative effectiveness of CBT, adaptive pacing or other interventions. Further, a critique of the Larun et al (2019) SRs is that there is not enough rigorous evidence of exercise therapy and CBT to help people with CFS (Larun et al., 2019; Vink and Vink-Niese, 2020). The critique made reference to included studies having: badly matched control groups; relying on an unreliable fatigue instrument as a primary outcome; outcome switching, P-hacking and ignoring evidence of harm (Vink and Vink-Niese, 2020). Electrical nerve stimulation, sleep and touch therapy, and behavioural self-management may be beneficial when; physical and psychological support is delivered in groups, people can plan their functional response to fatigue, strengthening rather than endurance is used to prevent deconditioning, fatigue is regarded in the context of an individual’s lifestyle, and home-based activities are used (Fowler-Davis 2021).

The NICE guidance for ME/CFS was originally published in 2007 and included recommendations for GET. However, the guidance was amended in 2021 (following expert consultation). The guidance now states that people experiencing fatigue ‘should undertake therapy options where they remain within their energy limits, and care should be given to undertake activities that do not worsen symptoms’ (National Institute for Health and Care Excellence, 2021). GET was removed from the guidance as ‘no evidence was identified for mortality, cognitive function, psychological status, pain, sleep quality, treatment-related adverse events, activity levels, care needs, and impact on families and carers. Therefore, these treatments may not be the preferred treatment for individuals living with Long COVID.

#### 3.1.3 Summary of primary studies

There are similarities between Long COVID and other conditions such as ME/CFS; however, the evidence base is limited. The primary studies included in this RR indicated that there should be a need-based focus on care and treatment for those with Long COVID in the same way care is presented to others with debilitating conditions (Lunt et al., 2022; Skilbeck, 2022; Wong et al., 2022). There is currently a lack of recent primary studies evaluating interventions to help people with post viral fatigue to return to normal activity, training and informal care.

### 3.2 Strengths and limitations of the available evidence

The strength of the evidence is weak. Ten studies were included in the review. Three systematic reviews were included, one of which focussed on Long COVID, one focussed on ME and CFS and one focussed on post viral fatigue syndrome (Chandan et al., 2022; Fowler-Davis et al., 2021; Larun et al., 2019). Four primary studies were included, three of which focussed on Long COVID (Lunt et al., 2022; Skilbeck, 2022; Wong et al., 2022) 2021) and one focussed on CFS (Nyland et al., 2014). There were also three relevant guidelines, two for Long COVID and one for ME/CFS. There was no evidence of interventions relating to return to training or returning to training, suggesting an evidence gap in these areas.

### 3.3 Implications for policy and practice

▪ Long COVID is still being established as a post-viral condition with many symptoms. The Welsh Government may seek to consider patient-centred treatment options such as occupational therapy, self-management therapy and talking therapy (such as Cognitive Behavioural Therapy) in the same way as for other similar conditions such as ME/CFS or other debilitating conditions.
▪ Return-to-work accommodations are needed for all workers unable to return to full-time employment.
▪ Long COVID is still being established as a post-viral condition with many symptoms. The Welsh Government may seek to consider patient-centred treatment options such as occupational therapy, self-management therapy and talking therapy (such as Cognitive Behavioural Therapy) in the same way as for other similar conditions such as ME/CFS or other debilitating conditions.
▪ Return-to-work accommodations are needed for all workers unable to return to full-time employment.

### 3.4 Strengths and limitations of this Rapid Review

#### 3.4.1 Strengths

- Some evidence included in this RR suggests that individuals with conditions similar to Long COVID can be supported back into work.
- Guidelines were also found that provide individuals with Long COVID and ME/CFS with information on how to self-manage their conditions.
- This review identified evidence which can help researchers prioritise future questions.

### 3.4.2 Limitations

- This review used a pragmatic search which will have limited the scope of the evidence found.
- Although there are similarities between Long COVID and other conditions such as ME/CFS, the evidence base is limited.
- The strength of the evidence is weak overall, and no extensive evidence base was identified.
- Only four identified papers were directly related to Long COVID.
- There was no evidence of interventions relating to returning to informal care, suggesting an evidence gap in this area.

## Data Availability

All data produced in the present study are available upon reasonable request to the authors

## Abbreviations

CBT: Cognitive Behavioural Therapy
CFS: Chronic Fatigue Syndrome
CIPD: Chartered Institute of Personnel and Development
COVID-19: Coronavirus disease 19
FDA: Food and Drug Administration
GET: Graded Exercise Therapy
GP: General Practitioner
ME: Myalgic encephalomyelitis
NHS: National Health Service
NICE: The National Institute for Health and Care Excellence
OECD: Organisation for Economic Co-operation and Development
PRISMA: Preferred Reporting Items for Systematic Reviews and Meta-Analyses
Qfever: Q fever is a disease caused by the bacteria Coxiella burnetii. The “Q” comes from “Query” fever, the name of the disease until its true cause was discovered in the 1930s.
RCT: Randomised Controlled Trial
RES: Rapid Evidence Summary
RR: Rapid Review
SR: Systematic Review
UK: United Kingdom
USA: United States of America
WCEC: Wales COVID-19 Evidence Centre
WHO: World Health Organisation
WHODAS: World Health Organization Disability Assessment Schedule 2.0 12L (WHODAS)

## 5. RAPID REVIEW METHODS

### 5.1 Eligibility criteria

The eligibility criteria for the review is presented in Table 5.1, and is based on the Population, Intervention, Comparison and Outcome (PICO) framework (Schardt et al., 2007). The Rapid Review stakeholders are interested in Long COVID and activities of normal life, including employment, informal caring and all kinds of activities of normal life. The outcomes of interest are: ‘work experience,’ ‘access to work,’ ‘return to work’ ‘employment,’ ‘unemployment,’ ‘work absenteeism,’ ‘work presenteeism,’ ‘unpaid work and ‘informal caring.’

**Table 5.1.** Eligibility criteria.

|  | **Inclusion criteria** | **Exclusion criteria** |
| --- | --- | --- |
| **Population** | Adults with Long COVID or similar post-viral conditions characterised by chronic fatigue | People without Long COVID or similar post-viral conditions |
| **Intervention/exposure** | Career advice<br>Cognitive Behavioural Therapy (CBT)<br>Exercise Therapy<br>Guidance (including therapy and counselling)<br>Neurological services<br>Occupational health<br>Physiotherapy<br>Rehabilitation | Studies not evaluating an intervention |
| **Counter intervention** | Usual care | Studies not evaluating usual care |
| **Outcome measures** | Absenteeism<br>Access<br>Activities<br>Caring<br>Employment<br>Life activities<br>Presenteeism<br>Return to work<br>Training for work<br>Unpaid work<br>Work<br>Work experience | Any outcome not related to normal activities of life |
| **Setting** | Any setting including health care settings or community/non health care setting | Any settings not supporting people to return to normal activities of life |
| **Context** | Support for people with Long COVID or other post-viral conditions | Any context not supporting people to return to normal activities of life |
| **Study design** | Qualitative studies<br>Quantitative studies<br>Mixed methods studies<br>Systematic Reviews<br>Rapid Reviews<br>Scoping Reviews | Any paper not reporting primary evidence e.g. not editorial, letters, position papers or protocols |
| **Countries** | OECD countries | Non-OECD countries |
| **Language of publication** | English, Welsh, or French | Publications not In English, Welsh, or French |
| **Publication date** | January 2012 to September 2022 | Papers published pre-January 2012. |
| **Publication type** | Published, pre-print, grey literature. | N/A |
| **Other factors** | N/A | N/A |

**Abbreviations:** OECD - Organisation for Economic Co-operation and Development

### 5.2 Search strategy and literature search

Relevant clinical guidelines and existing systematic reviews of non-pharmaceutical interventions for improving function or activities of daily living in patients with post-vial syndromes or Long COVID were identified as part of a preliminary review of the literature. As the evidence base for primary studies was likely to be limited and to work within the scope of a rapid review, a pragmatic search strategy was developed. The search strategy was developed by the research team in consultation with an information scientist and can be seen in Appendix 1.

Key databases will be searched for studies published between January 2012 and September 2022. The date limitations will be applied to keep the search within the scope of a Rapid Review. Databases to be searched are:

Medline

ASSIA

PsycINFO

CINAHL

EmBASE

Cochrane Library

### 5.3 Study selection process

Database searches were conducted by two members of the core Rapid Review team from BIHMR. Using the Covidence data screening and data extraction software tool for systematic reviews, citations were screened on title and abstract by at least two members of the core Rapid Review team. Full-text articles were retrieved and further assessed for inclusion. Any queries regarding inclusion/exclusion were resolved by discussion between members of the review team.

Due to the time constraints of a Rapid Review, full double screening was not possible. However, a sample of citations was double screened by the review lead to ensure adherence to inclusion/exclusion criteria.

### 5.4 Data extraction

The data was extracted from the included studies using a pre-defined data extraction tool developed to capture all relevant data. Extracted data included study details such as author, year, setting, aim, design, population, sample size, type of study, method of analysis, key findings, and author conclusions.

Included papers were distributed among the six core members of the review team for data extraction. A sample of extracted studies was checked against the papers for accuracy by the review lead (LHS). A proportion of the papers (10%) were double extracted to check for discrepancies between reviewers.

### 5.5 Quality appraisal

Quality appraisals were carried out by members of the review team using the JBI critical appraisal tools. This includes the JBI case reports checklist (Munn et al., 2021), and the JBI cohort studies checklist (Moola et al., 2017) amongst others.

Members of the review team chose the most appropriate JBI critical appraisal tool. A quarter of critical appraisals were checked by a second reviewer. Discrepancies arising during the critical appraisal process were discussed until the review team reached an agreement. The quality appraisal Tables 6.3.1 – 6.3.5 are in Section 6.3 of this report.

### 5.6 Synthesis

A narrative synthesis (Popay et al., 2006) was used as an overarching analysis framework and was used in conjunction with other descriptive/statistical analysis to develop a textual summary of the selected studies. This method relies on the use of words and text to summarise and explain the findings and tell the story of the findings from the included studies (See the results in Section 2 of this Rapid Review).

## 6. EVIDENCE

### 6.1 Evidence selection flow chart

The evidence selection flowchart is shown in Figure 1.

**Figure 1.**
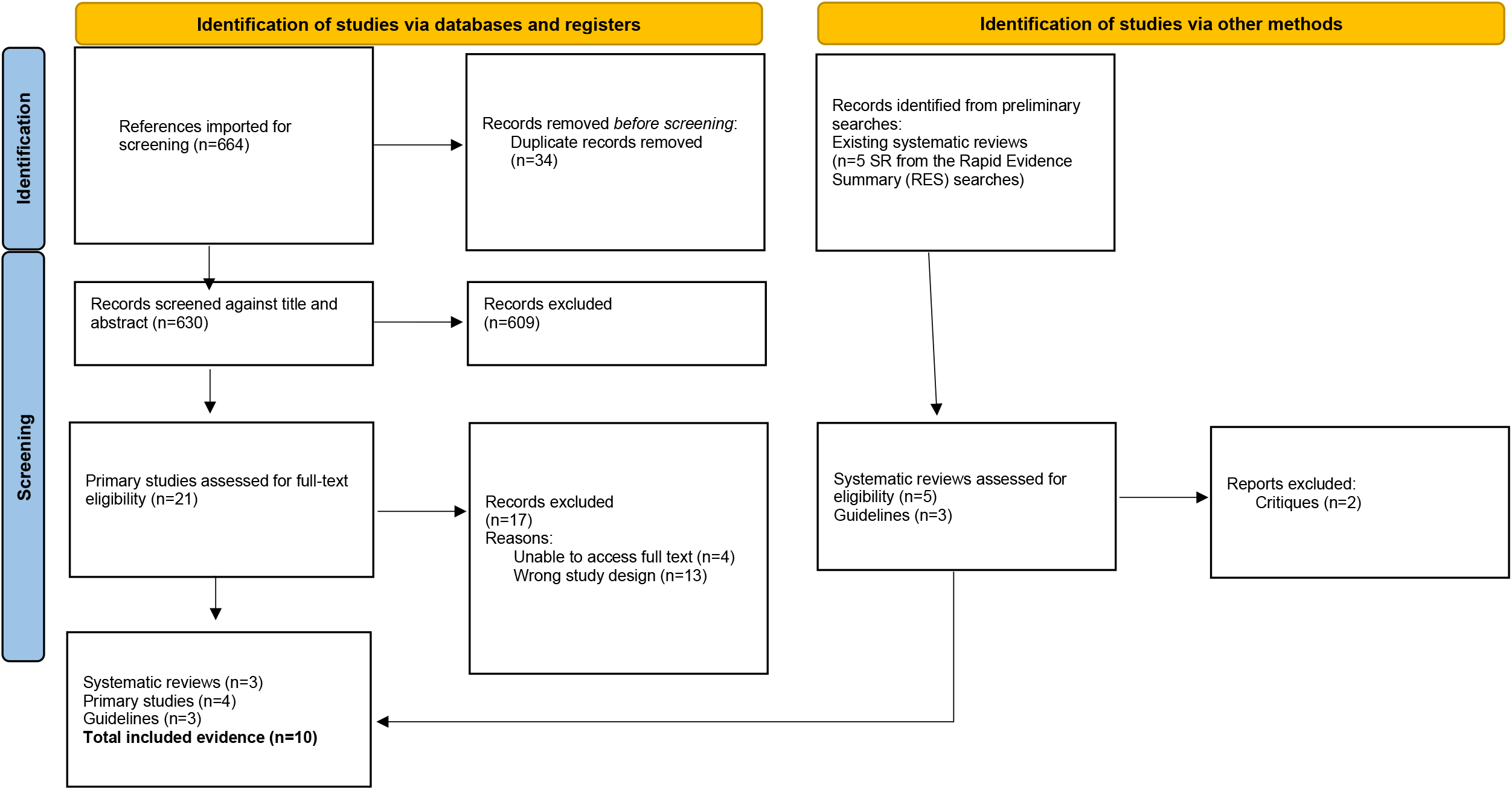
PRISMA study selection flowchart (Page et al., 2021)

### 6.2 Data extraction tables

Data extraction tables are presented in the results section (see Tables 2.1 and 2.2). Table 5.1 was the eligibility criteria and Table 6.1 is a map of the included evidence.

**Table 6.1.** Map of the support for Long COVID and similar post-viral syndromes.

| Type of evidence | Support for Long COVID or other post-viral syndromes |  |  |  |
| --- | --- | --- | --- | --- |
|  | Long COVID | ME/CFS | Other post-viral syndromes | Total |
| Systematic Review (SR) | Chandan et al (2022) | Larun et al (2019) | Fowler-Davis et al (2021) | 3 |
| Cohort study | Lunt et al (2022) | Nyland et al (2014) |  | 2 |
| Qualitative study | Wong et al (2022) |  |  | 1 |
| Case study | Skilbeck (2022) |  |  | 1 |
| Guides | Chartered Institute of Personnel and Development (2022) |  |  | 1 |
| Guidelines | Covid19: Rapid Guideline Managing the Longterm Effects of Covid19 (NICE, 2022) | NICE Guidelines NG206 (October 2021) |  | 2 |
| **Total** | **6** | **3** | **1** | **10** |

### 6.3 Quality appraisal tables

Members of the core review team chose the most appropriate JBI critical appraisal tool. A quarter of critical appraisals will be checked by a second reviewer. Discrepancies arising during the critical appraisal process will be discussed until an agreement is reached by the review team. When possible, studies will be graded as ‘very low’, ‘low’, ‘moderate’ or ‘high’ quality (See Table 6.3.1 to 6.3.5 below).

**Table 6.3.1.** JBI Critical analytical qualitative checklist (Joanna Briggs Institute, 2017a)

| Citation | Q1 Is there congruity between the stated philosophical perspective and the research methodology? | Q2 Is there congruity between the research methodology and the research question or objectives? | Q3 Is there congruity between the research methodology and the methods used to collect data? | Q4 Is there congruity between the research methodology and the representation and analysis of data? | Q5 Is there congruity between the research methodology and the interpretation of results? | Q6 Is there a statement locating the researcher culturally or theoretically? | Q7 Is the influence of the researcher on the research, and vice-versa, addressed? | Q8 Are participants, and their voices, adequately represented? | Q9 Is the research ethical according to current criteria or, for recent studies, and is there evidence of ethical approval by an appropriate body? | Q10 Do the conclusions drawn in the research report flow from the analysis, or interpretation, of the data? |
| --- | --- | --- | --- | --- | --- | --- | --- | --- | --- | --- |
| Wong et al (2021) | Yes | Yes | Yes | Yes | Yes | Yes | No | Yes | Yes | Yes |

**Table 6.3.2.** JBI Critical appraisal checklist for case reports (Gagnier et al., 2013)

| Citation | Q1. Were patient's demographic characteristics clearly described? | Q2. Was the patient's history clearly described and presented as a timeline? | Q3. Was the current clinical condition of the patient on presentation clearly described? | Q4. Were diagnostic tests or assessment methods and the results clearly described? | Q5. Was the intervention(s) or treatment procedure(s) clearly described? | Q6. Was the post-intervention clinical condition clearly described? | Q7. Were adverse events (harms) or unanticipated events identified and described? | Q8. Does the case report provide takeaway lessons? |
| --- | --- | --- | --- | --- | --- | --- | --- | --- |
| Skilbeck (2022) | Yes | Yes | Yes | Yes | Yes | Yes | No | Yes |

**Table 6.3.3.** JBI cohort checklist (Moola et al., 2017)

| Citation | Q1. Were the two groups similar and recruited from the same population? | Q2. Were the exposures measured similarly to assign people to both exposed and unexposed groups? | Q3. Was the exposure measured in a valid and reliable way? | Q4. Were confounding factors identified? | Q5. Were strategies to deal with confounding factors stated? | Q6. Were the groups/ participants free of the outcome at the start of the study (or at the moment of exposure)? | Q7. Were the outcomes measured in a valid and reliable way? | Q8. Was the follow up time reported and sufficient to be long enough for outcomes to occur? | Q9. Was follow up complete, and if not, were the reasons to loss to follow up described and explored? | Q10. Were strategies to address incomplete follow up utilized? | Q11. Was appropriate statistical analysis used? |
| --- | --- | --- | --- | --- | --- | --- | --- | --- | --- | --- | --- |
| Nyland et al 2014 | N/A | N/A | Yes | No | No | Yes | Yes | Yes | Yes | No | Yes |

**Table 6.3.4.** JBI Systematic Reviews and Research Syntheses Checklist (Joanna Briggs Institute, 2017b)

| Citation | Q1. Is the review question clearly and explicitly stated? | Q2. Were the inclusion criteria appropriate for the review question? | Q3. Was the search strategy appropriate? | Q4. Were the sources and resources used to search for studies adequate? | Q5. Were the criteria for appraising studies appropriate? | Q6. Was critical appraisal conducted by two or more reviewers independently? | Q7. Were there methods to minimize errors in data extraction? | Q8. Were the methods used to combine studies appropriate? | Q9. Was the likelihood of publication bias assessed? | Q10. Were recommendations for policy and/or practice supported by the reported data? | Q11. Were the specific directives for new research appropriate? |
| --- | --- | --- | --- | --- | --- | --- | --- | --- | --- | --- | --- |
| Fowler-Davis et al 2021 | Yes | Yes | Yes | Yes | Yes | Yes | Yes | Yes | Yes | Yes | Yes |
| Larun et al 2019 | Yes | Yes | Yes | Yes | Yes | Yes | Yes | Yes | Yes | Yes | Yes |
| Chandan et al 2022 | Yes | Yes | Unclear | Yes | Yes | Yes | Yes | Yes | Yes | Yes | Yes |

**Table 6.3.5.** MMAT Critical Appraisal of mixed methods studies (Munn et al., 2021)

| Study | MMAT<br>Mixed methods items |  |  |  |  | Score |
| --- | --- | --- | --- | --- | --- | --- |
|  | 5.1 | 5.2 | 5.3 | 5.4 | 5.5 |  |
|  | Is there an adequate rationale for using a mixed methods design to address the research question? | Are the different components of the study effectively integrated to answer the research question? | Are the outputs of the integration of qualitative and quantitative components adequately interpreted? | Are divergences and inconsistencies between quantitative and qualitative results adequately addressed? | Do the different components of the study adhere to the quality criteria of each tradition of the methods involved? |  |
| Lunt et al 2022 | Yes | Yes | Yes | Yes | Yes | High |

### 6.4 Information available on request

The RR protocol and list of excluded studies can be provided upon request to the lead corresponding author.

## 7.2 Acknowledgements

The authors would like to thank expert stakeholders including: Tracey Williams, Welsh Government; Rachel Wallbank, Cardiff and Vale University Health Board; Alexandra Strong and Nigel Pearson, Public Involvement Representatives.

Many thanks to Professor Adrian Edwards, Dr Ruth Lewis and Dr Alison Cooper (Wales COVID-19 Evidence Centre) for providing support to the review team: Elizabeth Gillen, Cardiff University, for reviewing our search strategy; Mr Mohammed Albustami and Dr Catherine Lawrence for lending their professional knowledge in ensuring the factual correctness of the details and for proofreading the report.

## 8. ABOUT THE WALES COVID-19 EVIDENCE CENTRE (WCEC)

The WCEC integrates with worldwide efforts to synthesise and mobilise knowledge from research.

We operate with a core team as part of Health and Care Research Wales, are hosted in the [Wales Centre for Primary and Emergency Care Research (PRIME)](http://www.primecentre.wales/), and are led by [Professor Adrian Edwards of Cardiff University](https://www.cardiff.ac.uk/people/view/123022-edwards-adrian).

The core team of the centre works closely with collaborating partners in [Health Technology Wales](https://www.healthtechnology.wales/), Wales Centre for Evidence-Based Care, [Specialist Unit for Review Evidence centre](https://www.cardiff.ac.uk/specialist-unit-for-review-evidence), [SAIL Databank](https://healthandcareresearchwales.org/about-research-community/secure-anonymised-information-linkage-sail-databank), Bangor Institute for Health & Medical Research/ Health and Care Economics Cymru, and the [Public Health Wales Observatory](https://phw.nhs.wales/services-and-teams/observatory/).

Together we aim to provide around 50 reviews per year, answering the priority questions for policy and practice in Wales as we meet the demands of the pandemic and its impacts.

### Director

Professor Adrian Edwards

### Website

https://healthandcareresearchwales.org/about-research-community/wales-covid-19-evidence-centre

## 9. APPENDIX

## APPENDIX 1: Search Strategy

### Support for Long COVID search strategy

#### Written for Medline and adapted for CINAHL, PsycINFO, ASSIA, Embase and the Cochrane Library

1. COVID-19/
2. Influenza, Human/
3. Epstein-Barr Virus Infection/
4. Meningitis/
5. Lyme Disease/
6. Q Fever/
7. Infectious Mononucleosis/
8. (Covid or* or COVID* or influenza or Epstein-Barr or Meningitis or Lyme disease or Q Fever or Infectious Mononucleosis or glandular fever),tw
9. 1 OR 2 OR 3 OR 4 OR 5 OR 6 OR 7 OR 8
10. (long or long term or long-term or persistent or chronic or recurring or recurrent or post viral or post-viral),tw
11. Employment/
12. Household Work/
13. Child Care/
14. Return to Work/
15. 11 OR 12 OR 13 OR 14
16. 9 AND 10 AND 15

## APPENDIX 2

**Resources searched during Rapid Review Searching**

APPENDIX 2: Resources searched during Rapid Review Searching
| Resource | Number of hits |
| --- | --- |
| Core non-COVID-19 specific resources |  |
| Medline/Embase/PsycInfo/ASSIA/CINAHL | 6 |
| Cochrane Library | 1 |
| Additional resources searched |  |
| Google Advanced Search | 1 |
| Google Scholar | 3 |
| **Total** | **11** |

## APPENDIX 3

**NIHR funded non-pharmacological studies for Long COVID (in progress)**

1. An activity tracking project (a Scotland based study) will track the physical activities of people with Long COVID to better manage their daily activity. End date: February 2023. Award ID: COV-LT2-0010
2. Long COVID Multidisciplinary Consortium (LOCOMOTION) (an England based study) will optimise Long COVID management across three settings of care (specialist Long COVID clinics; homes and self-management; primary care). End date: August 2023. Award ID: COV-LT2-0016
3. The LISTEN project (a UK study) will co-produce a package of support for individuals with Long COVID. This will include an economic analysis. End date: August 2023. Award ID: COV-LT2-0009
4. STIMULATE ICP Study (an England based study) will examine existing integrated care packages and design and test the best way of caring for people with Long COVID. End date: August 2023. Award ID: COV-LT2-0043
5. Long COVID Multidisciplinary Consortium (LOCOMOTION) (an England based study) will optimise Long COVID management across three settings of care (specialist Long COVID clinics; homes and self-management; primary care). End date: August 2023. Award ID: COV-LT2-0016
6. The Remote Diet Interventions to Reduce Long COVID Symptoms (ReDIRECT) (a Scotland based study) will investigate dietary intake and weight management of people with Long COVID to reduce symptoms such as fatigue, breathlessness and pain. End date: October 2023. Award ID: COV-LT2-0059
7. STIMULATE ICP Study (an England based study) will examine existing integrated care packages and design and test the best way of caring for people with Long COVID. End date: August 2023. Award ID: COV-LT2-0043

## REFERENCES

Amdal, C.D., Pe, M., Falk, R.S., Piccinin, C., Bottomley, A., Arraras, J.I., Darlington, A.S., Hofsø, K., Holzner, B., Jørgensen, N.M.H., Kulis, D., Rimehaug, S.A., Singer, S., Taylor, K., Wheelwright, S., Bjordal, K., 2021. Health-related quality of life issues, including symptoms, in patients with active COVID-19 or post COVID-19; a systematic literature review. Qual. Life Res. 30, 3367–3381.

Castro-Marrero, J., Faro, M., Zaragozá, M.C., Aliste, L., De Sevilla, T.F., Alegre, J., 2019. Unemployment and work disability in individuals with chronic fatigue syndrome/myalgic encephalomyelitis: A community-based cross-sectional study from Spain. BMC Public Health 19, 1–13.

Chandan, J.S., Brown, K., Simms-Williams, N., Camaradou, J., Bashir, N., Heining, D., Aiyegbusi, O.L., Turner, G., Cruz Rivera, S., Hotham, R., Nirantharakumar, K., Sivan, M., Khunti, K., Raindi, D., Marwaha, S., Hughes, S.E., McMullan, C., Calvert, M., Haroon, S., 2022. Non-pharmacological therapies for postviral syndromes, including Long COVID: a systematic review and meta-analysis protocol. BMJ Open 12.

Chartered Institute of Personnel and Development, 2022. Working with Long COVID: Guide for people professionals to provide support to those with Long COVID.

de Oliveira Almeida, K., Nogueira Alves, I.G., de Queiroz, R.S., de Castro, M.R., Gomes, V.A., Santos Fontoura, F.C., Brites, C., Neto, M.G., 2022. A systematic review on physical function, activities of daily living and health-related quality of life in COVID-19 survivors. Chronic Illn. 174239532210893.

Edwards, R.T., Spencer, L.H., Anthony, B., Bryning, L., 2019. Wellness in work: The economic arguments for investing in the health and wellbeing of the workforce in Wales [WWW Document]. URL https://cheme.bangor.ac.uk/documents/Wellness-in-Work-Report.pdf

Fowler-Davis, S., Platts, K., Thelwell, M., Woodward, A., Harrop, D., 2021. A mixed-methods systematic review of postviral fatigue interventions: Are there lessons for Long COVID? PLoS One 16, 1–23.

Gagnier, J.J., Kienle, G., Altman, D.G., Moher, D., Sox, H., Riley, D., 2013. Joanna Briggs Institute Checklist for Case Reports. JBI Crit. Apprais. Checkl. Case Reports 53, 1541–1547.

Gualano, M.., Rossi, M.., Borrelli, I., Santoro, P.., Amantea, C., Daniele, A., Tumminello, A., Moscato, U., 2022. Returning to work and the impact of post COVID-19 condition: A systematic review. Work.

Joanna Briggs Institute, 2017a. Checklist for Qualitative Research. Joanna Briggs Inst. 6.

Joanna Briggs Institute, 2017b. Checklist for Systematic Reviews and Research Syntheses. Joanna Briggs Inst.

Kenny, G., McCann, K., O’Brien, C., Savinelli, S., Tinago, W., Yousif, O., Lambert, J.S., O’Broin, C., Feeney, E.R., De Barra, E., Doran, P., Mallon, P.W.G., 2022. Identification of Distinct Long COVID Clinical Phenotypes Through Cluster Analysis of Self-Reported Symptoms. Open Forum Infect. Dis. 9.

Larun, L., Brurberg, K.G., Odgaard-Jensen, J., Price, J.R., 2019. Exercise therapy for chronic fatigue syndrome. Cochrane Database Syst. Rev. 2019.

Lunt, J., Hemming, S., Elander, J., Baraniak, A., Burton, K., Ellington, D., 2022. Experiences of workers with post-COVID-19 symptoms can signpost suitable workplace accommodations. Int. J. Work. Heal. Manag. 15, 359–374.

Moola, S., Munn, Z., Tufanaru, C., Aromataris, E., Sears, K., Sfetcu, R., Currie, M., Qureshi, R., Mattis, P., Lisy, K., Mu, P.-F., 2017. Checklist for Cohort Studies. Joanna Briggs Inst. Rev. Man. 1–7.

Munn, Z., Barker, T., Moola, S., Tufanaru, C., Stern, C., McArthur, A., Stephenson, M., Aromataris, E., 2021. Methodological quality of case series studies [WWW Document]. JBI Evid. Synth. URL https://jbi.global/critical-appraisal-tools (accessed 6.30.21).

National Institute for Health and Care Excellence, 2021. Myalgic encephalomyelitis (or encephalopathy)/chronic fatigue syndrome: diagnosis and management, NICE Guidelines.

National Institute for Health and Care Excellence (NICE), 2022. NICE Webpage [WWW Document]. URL https://www.nice.org.uk/

National Institute for Health and Care Excellence, Royal College of General Practitioners, Health Improvement Scotland, SIGN, 2022. Covid19: Rapid Guideline Managing the Longterm Effects of Covid19. 1.14 Publ. 01.03.2022 1–106.

NHS Plus, 2006. Occupational Aspects of the Management of Chronic Fatigue Syndrome: a National Guideline.

Nyland, M., Naess, H., Birkeland, J.S., Nyland, H., 2014. Longitudinal follow-up of employment status in patients with chronic fatigue syndrome after mononucleosis. BMJ Open 4, 1–7.

Popay, J., Roberts, H., Sowden, A., Petticrew, M., Arai, L., Rodgers, M., Britten, N., Roen, K., Duffy, S., 2006. Guidance on the Conduct of Narrative Synthesis in Systematic Reviews: A product from the ESRC Methods Programme [WWW Document]. URL https://www.lancaster.ac.uk/media/lancaster-university/content-assets/documents/fhm/dhr/chir/NSsynthesisguidanceVersion1-April2006.pdf

Ross, S.., Estok, R.., Frame, D., Stone, L.., Ludensky, V., Levine, C.., 2004. Disability and Chronic Fatigue Syndrome: A Focus on Function. Disabil. Chronic Fatigue Syndr. 164, 1089–1107.

Schardt, C., Adams, M.B., Owens, T., Keitz, S., Fontelo, P., 2007. Utilization of the PICO framework to improve searching PubMed for clinical questions. BMC Med. Inform. Decis. Mak. 7, 1–6.

Skilbeck, L., 2022. Patient-led integrated cognitive behavioural therapy for management of Long COVID with comorbid depression and anxiety in primary care - A case study. Chronic Illn. 18, 691–701.

UK Legislation, 2010. Equality Act 2010 [WWW Document]. URL https://www.legislation.gov.uk/ukpga/2010/15/contents (accessed 8.23.22).

Vink, M., Vink-Niese, F., 2020. Graded exercise therapy does not restore the ability to work in ME/CFS - Rethinking of a Cochrane review. Work 66, 283–308.

Welsh Government, 2017a. Prosperity for all: economic action plan [WWW Document]. URL https://gov.wales/sites/default/files/publications/2019-02/prosperity-for-all-economic-action-plan.pdf

Welsh Government, 2017b. Prosperity for All: The national strategy [WWW Document]. URL https://gov.wales/sites/default/files/publications/2017-10/prosperity-for-all-the-national-strategy.pdf

Welsh Government, 2021. Adferiad (Recovery) Long COVID programme [WWW Document]. URL https://gov.wales/adferiad-recovery-long-covid-programme-html x(accessed 8.22.22).

Wong, J., Kudla, A., Pham, T., Ezeife, N., Crown, D., Capraro, P., Trierweiler, R., Tomazin, S., Heinemann, A.W., 2022. Lessons Learned by Rehabilitation Counselors and Physicians in Services to COVID-19 Long-Haulers: A Qualitative Study. Rehabil. Couns. Bull. 66, 25–35.

Wong, T.L., Weitzer, D.J., 2021. Long COVID and myalgic encephalomyelitis/chronic fatigue syndrome (ME/CFS)-A systemic review and comparison of clinical presentation and symptomatology. Med. 57.

Zahra, D., Qureshi, A., Henley, W., Taylor, R., Quinn, C., Hardy, G., Newbold, A., Byng, R., 2014. The work and social adjustment scale: Reliability, sensitivity and value. Int. J. Psychiatry Clin. Pract. 18, 131–138.

